# Large language models outperform traditional natural language processing methods in extracting patient-reported outcomes in IBD

**DOI:** 10.1101/2024.09.05.24313139

**Authors:** Perseus V. Patel, Conner Davis, Amariel Ralbovsky, Daniel Tinoco, Christopher Y.K. Williams, Shadera Slatter, Behzad Naderalvojoud, Michael J. Rosen, Tina Hernandez-Boussard, Vivek Rudrapatna

## Abstract

**Background and Aims:** Patient-reported outcomes (PROs) are vital in assessing disease activity and treatment outcomes in inflammatory bowel disease (IBD). However, manual extraction of these PROs from the free-text of clinical notes is burdensome. We aimed to improve data curation from free-text information in the electronic health record, making it more available for research and quality improvement. This study aimed to compare traditional natural language processing (tNLP) and large language models (LLMs) in extracting three IBD PROs (abdominal pain, diarrhea, fecal blood) from clinical notes across two institutions.

**Methods:** Clinic notes were annotated for each PRO using preset protocols. Models were developed and internally tested at the University of California San Francisco (UCSF), and then externally validated at Stanford University. We compared tNLP and LLM-based models on accuracy, sensitivity, specificity, positive and negative predictive value. Additionally, we conducted fairness and error assessments.

**Results:** Inter-rater reliability between annotators was >90%. On the UCSF test set (n=50), the top-performing tNLP models showcased accuracies of 92% (abdominal pain), 82% (diarrhea) and 80% (fecal blood), comparable to GPT-4, which was 96%, 88%, and 90% accurate, respectively. On external validation at Stanford (n=250), tNLP models failed to generalize (61-62% accuracy) while GPT-4 maintained accuracies >90%. PaLM-2 and GPT-4 showed similar performance. No biases were detected based on demographics or diagnosis.

**Conclusions:** LLMs are accurate and generalizable methods for extracting PROs. They maintain excellent accuracy across institutions, despite heterogeneity in note templates and authors. Widespread adoption of such tools has the potential to enhance IBD research and patient care.

## Introduction

Patient-reported outcomes (PROs) provide key information on disease activity in randomized trials, treatment guidelines, and clinical practice^1–4^. PROs are commonly documented as free-text data in electronic health records (EHRs). Manual extraction of PROs for research is labor-intensive, difficult to sustain, and prone to human error ^5^. Due to the historical absence of computational methods for querying these data, many IBD studies using EHRs have excluded PROs and other information found in clinical notes, increasing the risk of bias.

Traditional natural language processing (tNLP), centered on rules-based approaches, can transform free-text into analysis-ready data, but suffers from variable accuracy and labor-intensiveness. Recently, large language models (LLMs), pre-trained to understand the contextual relationships in language, are showing promise in curating clinical information^6,7^. However, their generalizability across medical centers remains unclear.

In this study, we compared the effectiveness of tNLP versus LLMs for extracting 3 IBD PROs (abdominal pain; diarrhea; fecal blood), as a first step to enable better research and quality improvement in IBD. Given the importance of such tools to maintain high performance across institutions, we externally validated our models at a second institution to test generalizability.

## Methods

To create the clinical note datasets, we queried EHR databases at the University of California, San Francisco (UCSF) and Stanford University to extract adult IBD clinic notes based on pre-set inclusion and exclusion criteria. These notes included adult patients with IBD seen by a nurse practitioner or physician at an IBD clinic between June 1, 2012, to February 1, 2022 (Table 1). Two physicians annotated note samples per a pre-set protocol (Supplemental methods). Discrepancies were addressed through discussion between the two annotators, and if needed a third gastroenterologist was consulted. Inter-annotator agreement scores were calculated for each symptom using 100 randomly selected notes from the UCSF training dataset.

**Table 1:**
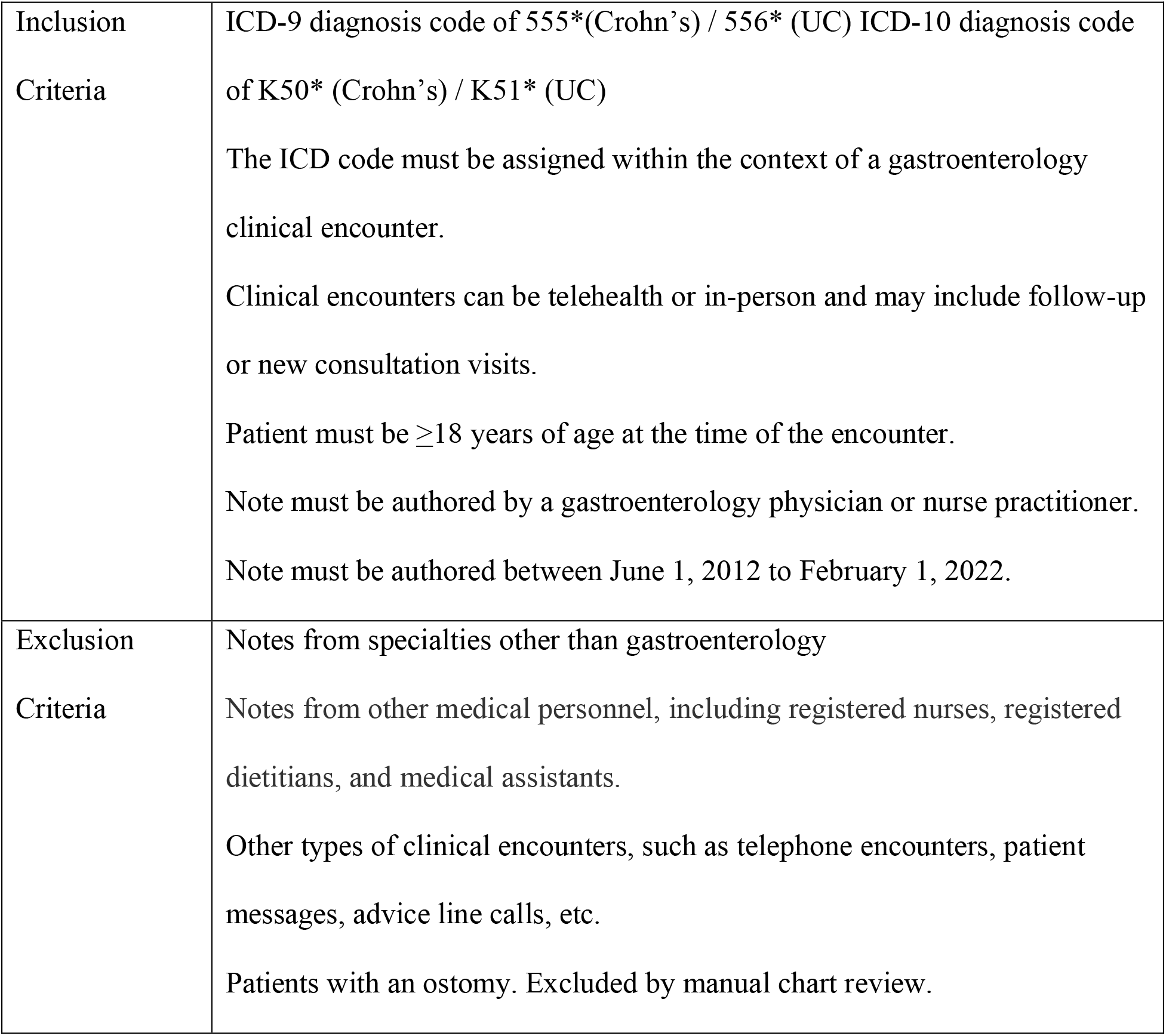
Inclusion and exclusion criteria for our cohorts at both institutions.

We observed that the original corpus of notes was “class imbalanced”, where some symptoms (e.g., abdominal pain) were much less commonly documented than the converse (e.g., no abdominal pain). This was notable because machine learning models learn best when the training dataset is “class balanced”, with a roughly equal number of positive and negative examples. To overcome this, we used MedSpaCy^8^, an open-source named entity recognition tool, to prescreen the overall note corpus and selectively identify notes corresponding to the underrepresented class. We then selectively annotated these notes to achieve greater class balance. This process resulted in slightly different sizes of training examples for each PRO, reflecting baseline differences in class balance across each symptom.

The UCSF dataset (n=879 notes) was used for both training and testing, with different train/test splits for each PRO (Figure 1). This dataset also underwent note preprocessing to isolate the history of present illness or interval history whenever available. By contrast, the Stanford dataset (n=250), extracted and annotated using the same protocol, was used for external validation only, with no preprocessing. This was done to assess the models’ inferential abilities without requiring institution-specific pre-processing based on specific note templates, therein reflecting a higher bar for generalizability.

**Figure 1:**
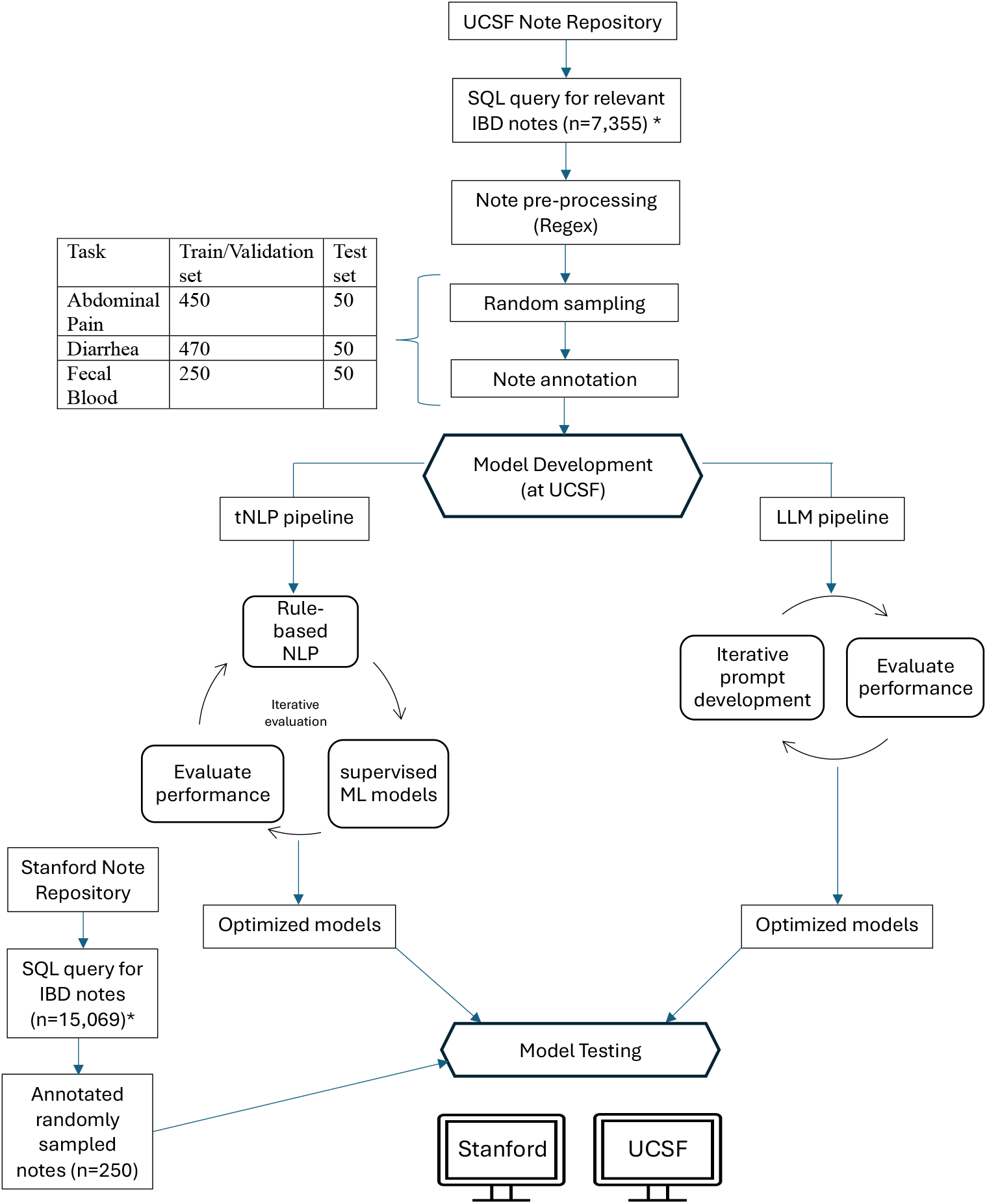
Methodology flowchart. *\*SQL query for IBD notes were based on inclusion and exclusion criteria (see Table 1). Annotation protocol, SQL codes, and final LLM prompts are publicly available at GitHub (see supplemental methods for URL)*

We built tNLP models that use expert-defined rules (e.g., symptom definitions, synonyms) and predictive features identified by supervised machine learning. See Supplemental Methods for full details of model development. Models were selected based on accuracy on the training data, then evaluated once on holdout UCSF and Stanford test sets without any subsequent modification of the model selection procedure (Figure 1).

We also evaluated two LLMs. Institutional governance permitted access to GPT-4 (UCSF, Stanford) and PaLM-2 (Stanford). We incorporated the annotation protocol into our prompts and used the UCSF training data for prompt engineering (Supplemental Methods). PaLM-2 used the same prompts designed for GPT-4.

At both institutions, we used holdout test sets to compare tNLP versus LLM performance. Our primary endpoint, accuracy, was the proportion of model-generated labels that matched the manual annotation of each PRO. We also assessed sensitivity, specificity, positive predictive value (PPV), and negative predictive value (NPV). To better understand the limitations of the LLM, we reviewed the LLM-provided reasoning for each incorrectly coded note.

We conducted a fairness assessment based on demographics and diagnosis. Binary variables were used for gender (male/female), race (white/non-white), and disease (Crohn’s Disease/Ulcerative Colitis). For race, the binarization into white/non-white allowed us to maintain statistical power given the diversity of races represented in our cohort. For age, we compared groups above and below the median age to maintain statistical power. We were unable to assess for differences in ethnicity as >95% of our cohort was non-Hispanic. In recognizing the limitations of our approach, we also randomly selected a set of 10 notes for each PRO where no mention of race was present, added a mention of race and ethnicity (White, African American, Asian, Hispanic, Non-Hispanic), and evaluated if the answer provided by the LLM changed amongst the different groups^9^. We were unable to assess differences in accuracy based on insurance status (public versus commercial) because only 8.4% (21/250) of the encounters included patients on public insurance.

The Human Research Protection Program Institutional Review Board at UCSF (IRB#18-24588) and Stanford University (IRB #47644) approved this study.

## Results

At UCSF, most notes included non-Hispanic (91.2%), white (71.2%), female (52.3%) patients with Crohn’s Disease (58.9%) and a median age of 37 (range: 18-86) years. At Stanford, most notes included non-Hispanic (95.2%), white (60%), male (58.4%) patients with Crohn’s Disease (55.2%) and a median age of 35 (range: 18-65) years.

Inter-annotator agreement was 91-93% for each PRO. In the UCSF test set (n=50), abdominal pain (AP), fecal blood (FB), and diarrhea (DH) were present in 28%, 26%, and 54% of the notes, respectively. In the Stanford test set (n=250), the PROs were present in 30.4% (AP), 25.4% (FB), and 44% (DH) of the notes.

In the UCSF test set, tNLP models showed similar performance to GPT-4. Specifically, they achieved accuracies of 92% (AP), 82% (DH) and 80% (FB); compared to 96% (AP), 88% (DH), and 90% (FB) as achieved by GPT-4. GPT-4 had 4-10% numeric superiority across PROs, but this was not statistically significant (Figure 2). The tNLP models showed poor generalizability—61-62% accuracy at Stanford—while GPT-4 maintained high performance across institutions. PaLM-2 and GPT-4 were equally accurate at Stanford (Figure 2). GPT-4 had better PPVs across PROs, but neither LLM was clearly superior (Table 2).

**Table 2:** Secondary Metrics for each model at both institutions with their associated 95% confidence intervals. The primary endpoint for each model was accuracy. Secondary endpoints included sensitivity, specificity, positive and negative predictive values. The ‘traditional’ natural language processing (NLP) model rule-based features and supervised learning models. Both large language models (GPT-4 and PaLM-2) were prompt-engineered using the same prompt.

| **Model** | **Institution** | **Task** | **Sensitivity** | **Specificity** | **Positive Predictive Value** | **Negative Predictive Value** |
| --- | --- | --- | --- | --- | --- | --- |
| Traditional NLP | UCSF (n=50) | Abdominal pain | 0.86<br>[0.76-0.96] | 0.93<br>[0.86-1.00] | 0.86<br>[0.76-0.96] | 0.94<br>[0.81-1.00] |
|  |  | Diarrhea | 0.85<br>[0.75-0.95] | 0.78<br>[0.67-0.89] | 0.82<br>[0.71-0.93] | 0.82<br>[0.71-0.93] |
|  |  | Fecal Blood | 0.79<br>[0.68-0.90] | 0.96<br>[0.91-1.00] | 0.86<br>[0.76-0.96] | 0.75<br>[0.63-0.87] |
|  | Stanford (n=250) | Abdominal pain | 0.28<br>[0.22-0.34] | 0.76<br>[0.71-0.81] | 0.33<br>[0.27-0.39] | 0.71<br>[0.65-0.77] |
|  |  | Diarrhea | 0.25<br>[0.20-0.30] | 0.89<br>[0.85-0.93] | 0.63<br>[0.57-0.69] | 0.61<br>[0.55-0.67] |
|  |  | Fecal Blood | 0.36<br>[0.30-0.42] | 0.71<br>[0.65-0.77] | 0.30<br>[0.24-0.36] | 0.75<br>[0.70-0.80] |
| GPT-4 | UCSF (n=50) | Abdominal pain | 0.93<br>[0.86-1.00] | 0.97<br>[0.92-1.00] | 0.93<br>[0.86-1.00] | 0.97<br>[0.92-1.00] |
|  |  | Diarrhea | 0.77<br>[0.65-0.89] | 1 | 1 | 0.80<br>[0.69-0.91] |
|  |  | Fecal Blood | 0.70 | 0.97 | 0.90 | 0.90 |
|  |  |  | [0.56-0.82] | [0.92-1.00] | [0.82-0.98] | [0.82-0.98] |
|  | Stanford<br>(n=250) | Abdominal<br>pain | 0.88<br>[0.84-0.92] | 0.98<br>[0.96-1.00] | 0.94<br>[0.91-0.97] | 0.95<br>[0.92-0.98] |
|  |  | Diarrhea | 0.80<br>[0.75-0.85] | 0.98<br>[0.96-1.00] | 0.97<br>[0.95-0.99] | 0.86<br>[0.82-0.90] |
|  |  | Fecal Blood | 0.91<br>[0.87-0.95] | 0.99<br>[0.98-1] | 0.97<br>[0.95-0.99] | 0.96<br>[0.94-0.98] |
| PaLM-2 | Stanford<br>(n=250) | Abdominal<br>pain | 0.96<br>[0.94-0.98] | 0.92<br>[0.89-0.95] | 0.84<br>[0.79-0.89] | 0.98<br>[0.96-1.00] |
|  |  | Diarrhea | 0.89<br>[0.85-0.93] | 0.88<br>[0.84-0.92] | 0.85<br>[0.81-0.89] | 0.91<br>[0.87-0.95] |
|  |  | Fecal Blood | 0.92<br>[0.89-0.95] | 0.96<br>[0.94-0.98] | 0.88<br>[0.84-0.92] | 0.97<br>[0.95-0.99] |

**Figure 2:**
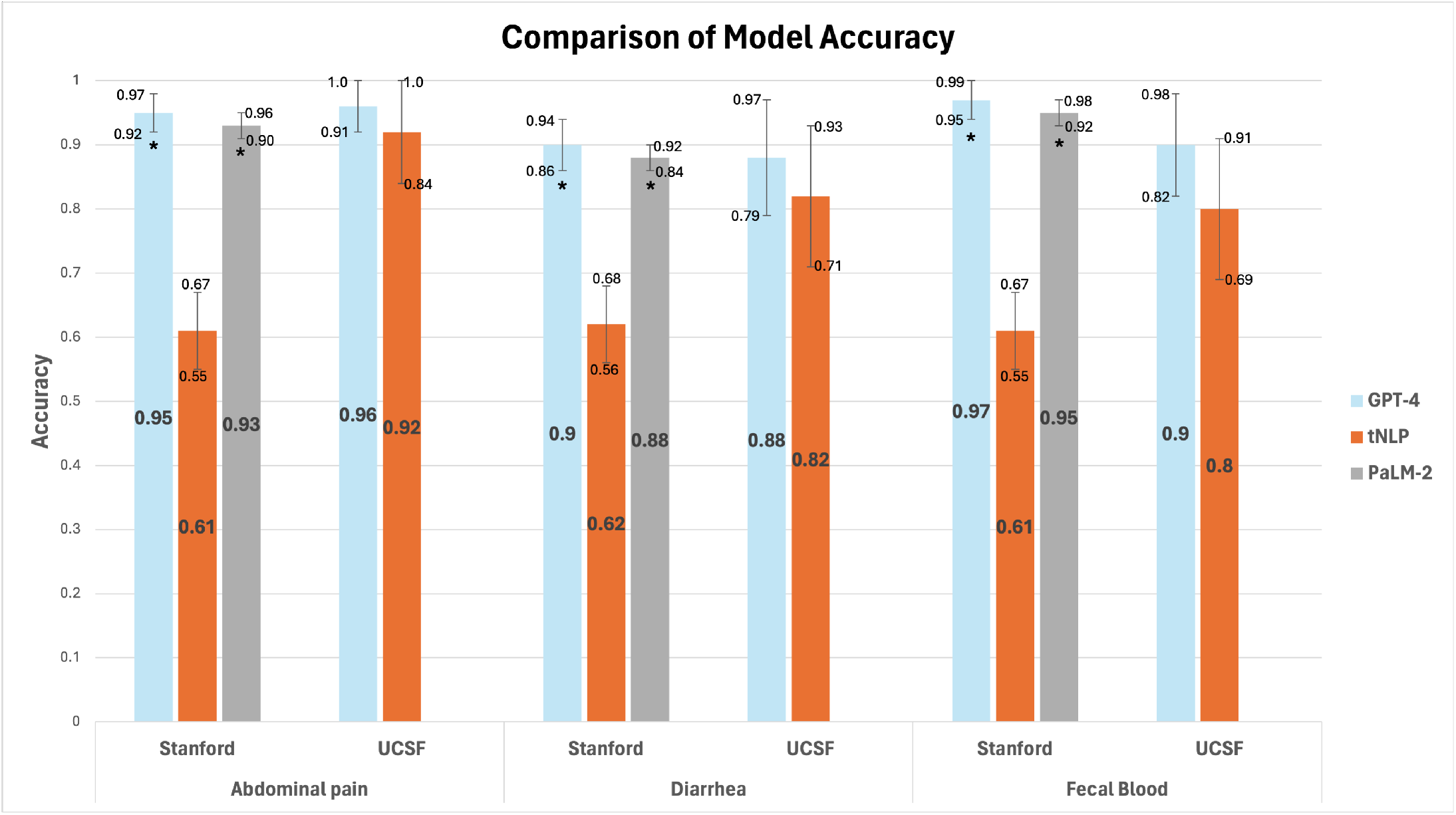
Bar graph comparing model accuracy of each model at both institutions.

To guide future model development and better understand shortcomings of our models, we conducted an error analysis on both LLMs as they were the best performing models on external validation. For abdominal pain, the majority of GPT-4’s errors misclassified symptom-positive notes as negatives. This was resultant from classifying improving symptoms, mild symptoms, or low symptom frequency as absent symptoms. In contrast, PaLM-2 misclassified abdominal pressure as pain, and assumed that the presence of other gastrointestinal symptoms indicated the likely presence of abdominal pain. Contradictory information, a consequence of note copy-forwarding, was a common source of error for both LLMs. For the fecal blood model, both LLMs misclassified minimal or intermittent hematochezia as the absence of fecal blood.

GPT-4 misclassified recurrent episodes (i.e.-“blood in stools 3x/week”) of blood as ‘no bleeding’, including when the last episode of blood was within the past week. On the other hand, PaLM-2 misclassified recently resolved episodes (i.e.-“fecal blood recently resolved over past 2 weeks”) as present. For the diarrhea model, both LLMs had similar errors. They struggled to accurately classify patients with wide ranges of stools consistency (“formed to loose stools”), bowel movement frequency (“1-5 stools/day”), and made arithmetic errors when calculating the total number of stools per day if providers wrote descriptors such as “2 stools in the morning and 3 stools in the evening”.

We performed our fairness analysis on clinic notes from Stanford University (n=250) because of the limited size of our UCSF test set (n=50). Results are shown in supplemental tables 1-8. We found no significant differences in performance across all model metrics for any of the subgroups analyzed. This was conducted on both LLMs (GPT-4 and PaLM-2) as they were the best performing models on external institution testing. Additionally, neither LLM changed any note classifications based on synthetically generated additions of race/ethnicity into the clinic note.

## Discussion

In this multicenter study, we compared traditional and state-of-the-art tools to extract IBD PROs from clinical notes. Traditional models achieve high institution-specific accuracy, and our open-source code provides a framework for building them. However, variations in documentation styles across institutions limited their generalizability. LLMs show resiliency across domain shifts; GPT-4 outperformed tNLP across institutions, in test sets that spanned 10-years, multiple authors, note templates, and changes in IBD guidelines. PaLM-2 and GPT-4 demonstrated similar accuracy. We anticipate that, due to their broad training corpus, these general-use LLMs will yield institution-agnostic results, but future studies are needed.

Furthermore, prompt-engineering LLMs was less labor-intensive. While the process of creating and testing tNLP pipelines spanned nearly 2 years, the LLMs were prompt-engineered in 3 months. These efficiencies will likely increase as baseline LLM performance improves. Our fairness analyses showed no LLM biases related to disease or demographics.

Strengths of this study include multiple model types, centers, thorough model assessment including fairness and errors, and use of a dataset with high inter-annotator agreement.

Additionally, the use of real-world data exposed the LLMs to issues such as copy-forwarding of notes. Despite this, Our study also shows the robustness of prompts across LLMs, further supporting the generalizability and utility of these models.

We acknowledge several limitations. We were unable to perform a multicenter assessment of PaLM-2 due to institutional limitations at UCSF. However, our analysis implies the comparability and generalizability of these models. Our fairness analysis was underpowered; additional testing is needed prior to widespread deployment. Lastly, our annotation definitions may not have universal agreement. While future protocols may differ, our findings suggest that LLMs are capable of handling varied definitions.

Overall, LLMs outperformed tNLP methods at accurately extracting IBD PROs from clinical notes across institutions. This multi-site study also yielded important insights for the field of clinical informatics as effective strategies for harmonizing unstructured data preclude high-quality, cost-effective studies on treatment outcomes. While this study was IBD-specific, the promising results support that these tools have the potential to open new areas of investigation across diseases. Future studies are needed to build upon our binary classification of PROs and characterize symptom severity across a gradient. In our error analysis, we found that that LLMs were more likely to mislabel mild or infrequent symptoms, but dedicated studies utilizing models focused on discerning severity are needed to validate these findings. Clinically, integrating LLM-based tools as decision-aids could streamline gastroenterology referrals to expedite diagnoses, and improve tracking and follow-up of symptomatic patients. Our works builds upon previous NLP work in IBD focused on extracting extraintestinal manifestations, classifying disease phenotype, and categorizing patient-physician interactions for future triage^10– 14^. Ultimately, improved monitoring using these tools can potentially improve time-to-treatment and limit disease sequelae. By employing LLM-based models across all notes throughout a patient’s disease trajectory, and linking the results with structured EHR data like laboratory values, future studies may utilize NLP tools to evaluate treatment outcomes, such as improvement in PROs following initiation of biologic therapy. Additionally, as LLM integration into medicine grows, developing similar tools is vital to bridging the digital divide to allow less-resourced institutions to participate in research consortia, and to utilize automated tools to optimize patient outcomes.

## Data Availability

All data produced in the present study are available upon reasonable request to the authors

https://github.com/rwelab/IBDNLP/tree/main

## Acknowledgements

The authors thank the Academic Research Services and Research Information Technology groups at UCSF, and Stanford Medicine Technology & Digital Solutions for enabling this work via HIPAA-compliant portals for GPT-4 and PaLM-2. PVP also thanks Stanford Children’s Health IBD and Celiac Center for their support of this work.

