## Supplemental Materials for "Large language models outperform traditional natural language processing methods in extracting patient-reported outcomes in IBD"

**Supplemental Materials Legend**

1. Supplemental Methods
   a. Clinical note annotation protocol
   b. Model development, training, and testing
2. Prompts used for both large language models
3. Protocol and code availability on GitHub
4. Data Security
5. Supplemental tables 1-8 for the fairness analyses based on demographics and disease diagnosis.

**Supplemental Methods**

*Clinical note annotation protocol*

Notes were categorized as “yes” [symptom is present], “no” [symptom is absent], “inferred yes” [symptom is likely present], “inferred no” [symptom is likely absent], or “missing” [no documentation regarding the presence/absence of a symptom]. For the purposes of streamlining the algorithm, this multi-stratum classification was simplified by combining “yes” with “inferred yes”, and “no” with “inferred no”, creating a binary classification schema. Intercoder reliability scores were calculated for classification of each note [yes/no] to establish a gold standard against which our algorithm was compared. For the symptom of “abdominal pain” we encountered a class imbalance in which roughly 80% of the notes were classified as positive. We used a prepackaged MedSpaCy pipeline to screen the note corpus for notes indicative of a patient without abdominal pain, and then manually annotated another 100 notes to achieve better balance for model training. Our annotation protocols were uploaded to GitHub (<https://github.com/rwelab/IBDNLP/tree/main>).

*Model development, training, and testing*

Both traditional natural language processing (tNLP) and large language models (LLMs) aim to understand and generate language. The traditional methods employed custom-built rules to define content-specific features like clinical remission, regular expressions to define a list of synonyms for symptoms, grammatical tense parsers to identify the present, and supervised machine learning that required content-specific manually annotated training data. On the other hand, LLMs such as GPT-4 and PaLM-2 are unsupervised, pre-trained models that generate a probabilistic output based on the task (prompt) described by a human.

For the tNLP models, we began by pre-processing notes using regular expression (RegEx) parsers to extract the History of Present Illness and/or Interval History sections from notes. We chose to prioritize data under these subheadings as they are most likely to contain the accurate, current information. If both headings were present, we prioritized information under the interval history section. These preprocessing pipelines are highly institution-specific as the RegEx code relies on upon start and end points for data extraction that are tailored to the note template used at that institution. For example, our interval history parser would have failed to work on follow-up clinic note templates that used a subheading of “interim history”, or in note templates where the order of the note sections (i.e.- interval history, medications, allergies, physical exam, etc.) differed. For institutions replicating our model, we recommend re-coding this area of the pipeline.

Following the pre-processing step, we then created the rules-based section of the symptom extraction pipeline using the key features replicating the criteria used by the human annotators (i.e.- consistency of stool consistency, bowel movement frequency, synonyms of pain, and anatomic descriptors that relate to the abdomen). We used our training data and clinical expertise to develop a comprehensive list of the ways providers may document or describe these symptoms within a clinical note. Bespoke functions were then developed for negation detection (i.e.- “patient denies blood in stool”), antonyms (i.e.- “patient is constipated”), specialized terms such as clinical remission. To address contradictory information that likely stemmed from copy-forwarding of previous notes, we implemented a frequency counter to assess the number of positive and negative mentions for a specific PRO. Once key words related to each symptom were identified, our pipeline identified the grammatical tense of the sentence in which the keyword was present to help delineate if the symptom was ongoing or occurred in the past.

Once all key features had been extracted from the note, models were trained on these extracted features, and then fine-tuned based on feature selection, hyperparameter tuning and feature engineering. We used Term-Frequency-Inverse-Document-Frequency (TFIDF) and SpaCy vectorization to establish a baseline performance to measure against and tested multiple machine learning models including but not limited to Random Forest, Support Vector Machine, Logistic Regression and Naïve Bayes for both the vectorizations and the data augmented by our pipeline. We also trialed Deep Learning models including continuous bag of words, long-short term memory, and a pretrained Bidirectional Encoder Representations from Transformers (BERT) models from HuggingFace for this process but found both to be sub-optimal for this task. Of the models that were tested, XG Boost had the highest accuracy for abdominal pain and fecal blood, and the lowest burden for fine-tuning and was chosen for use in our model testing. Logistic Regression model was selected for diarrhea. We performed further hyperparameter tuning and executed a 10-fold cross validation over a random grid hyperparameter search using 90% of the training dataset to assess the stability of the model, with the remaining 10% to finalize the hyperparameter selection. Subsequently, each model was tested on the holdout test set at UCSF (50 notes each), and no future model adjustments were made. The same models that were used in the UCSF test set were subsequently used for external testing at Stanford.

**Large Language Model Prompts**

The LLM prompts were developed using the rules given to human annotators. We used notes from the training datasets at UCSF to create and develop these prompts. As part of the ‘task’ section of the prompt, we had GPT-4 provide it’s reasoning for the classification. During prompt development, we used this as feedback for the incorrectly labeled notes, and adjusted the prompt when we noticed a pattern in incorrect labels. Once prompts were achieving ≥ 90% accuracy on training data, we deemed them to be finalized. No further prompt adjustments were made. These finalized prompts were then tested on a hold-out test set at UCSF (n=50 notes), and notes at Stanford (n=250 notes).

**Abdominal Pain**

You are an expert gastroenterologist tasked to identify the presence of current abdominal pain based on the clinical notes of patients with inflammatory bowel disease. Below is the clinical note for one patient:
###NOTE START###
XXXXX

###NOTE END###
Below are the instructions to determine, using only information in the above clinical note, whether the patient has current abdominal pain.
###INSTRUCTIONS START###
Definition of current abdominal pain:
- Any synonyms for pain, including 'pain', 'discomfort' or 'cramping'. The sensation of 'pressure' (e.g abdominal pressure) does not count as pain and can be ignored (assuming no other synonyms of pain are present).
- EITHER symptoms must be referred to in the present tense OR symptoms must have been present AT ANY TIME within the past week (7 days) to be classed as CURRENT abdominal pain (even if the patient has no abdominal pain at time of visit).
- References to 'pain' or 'discomfort' must be associated with an anatomic location relation to the abdomen. Common anatomic locations related to the abdomen include, but are not limited to:
-- Epigastric; Periumbilical; Right upper quadrant (RUQ); Right lower quadrant (RLQ); Left upper quadrant (LUQ); Left lower quadrant (LLQ); Right sided; Left sided; Lower abdomen; Upper abdomen; Diffuse or generalized pain across the abdomen; Pelvic; Stomach; Abd/Abdo; Flank
- 'Cramping' does not need to have an anatomic identifier associated with it to count as abdominal pain - i.e. any mention of 'cramping' qualifies as abdominal pain.
Edge cases:
- 'Clinical remission' - if the note states that a patient is in remission, then assume abdominal pain is NOT present unless there is any other information in the note to suggest the presence of current abdominal pain.
- 'Asymptomatic' or 'doing well' - if a note does not specifically mention the presence of abdominal pain, but states the patient is asymptomatic or doing well, assume abdominal pain is NOT present.
- If other gastrointestinal symptoms are documented in the note, but there is no mention of abdominal pain, assume that abdominal pain is NOT present.
- Contradictory information: some notes may contain contradictory information as to the presence/absence of abdominal pain. If this is the case, use clinical judgement to decide whether the patient has current abdominal pain as defined above.
###INSTRUCTIONS END###
Task: Use the above clinical note and the accompanying instructions to determine whether the patient has current abdominal pain. Give an explanation and return one of the following labels: {'0: No current abdominal pain', '1: Current abdominal pain'}

**Fecal Blood**

You are an expert gastroenterologist tasked to identify the presence of current fecal blood based on the clinical notes of patients with inflammatory bowel disease. Below is the clinical note for one patient:
###NOTE START###
xxxxxx
###NOTE END###
Below are the instructions to determine, using only information in the above clinical note, whether the patient has current fecal blood.
###INSTRUCTIONS START###
Definition of current fecal blood:
- Any synonyms of fecal blood including: 'fecal blood', 'bloody stools', 'bloody BMs', 'rectal bleeding' 'stools/BMs with blood', 'hematochezia', 'melena', 'tarry stools', 'bright red blood/BRB', 'bright red blood per rectum/BRBPR', 'rectal bleeding', '+ blood in stool', 'blood in toilet', 'blood-streaked stools', 'flecks of blood in stool', AND
- EITHER symptoms must be referred to in the present tense OR symptoms must be present within the past week (7 days) to count as CURRENT fecal blood.
Edge cases:
- 'Clinical remission' - if the note states that a patient is in remission, then assume fecal blood is NOT present unless there is any other information in the note to suggest the presence of current fecal blood.
- 'Asymptomatic' or 'doing well' - if a note does not specifically mention the presence of fecal blood, but states the patient is asymptomatic or doing well, assume fecal blood is NOT present.
- If other gastrointestinal symptoms are documented in the note, but there is no mention of fecal blood, assume that fecal blood is NOT present.
- Contradictory information: some notes may contain contradictory information as to the presence/absence of fecal blood. If this is the case, use clinical judgement to decide whether the patient has current fecal blood as defined above.
###INSTRUCTIONS END###
Task: Use the above clinical note and the accompanying instructions to determine whether the patient has current fecal blood. Give an explanation and return one of the following labels: {'0: No current fecal blood', '1: Current fecal blood'}

**Diarrhea**

You are an expert gastroenterologist tasked to identify the presence of current diarrhea based on the clinical notes of patients with inflammatory bowel disease. Below is the clinical note for one patient:
###NOTE START###
xxxxxxx
###NOTE END###
Below are the instructions to determine, using only information in the above clinical note, whether the patient has current diarrhea.
###INSTRUCTIONS START###
Definition of current diarrhea - ANY one of the following three criteria must be present within the past week (7 days) to count as CURRENT diarrhea:
a) Bristol stool scale score of 5, 6, or 7, OR
b) More than 3 bowel movements per day (per 24 hour period, including day and night-time bowel movements) REGARDLESS OF CONSISTENCY OF STOOL (if a range is given, use the higher number in the range to determine if patient has more than 3 bowel movements per day [e.g: if a range of 2-5 bowel movements is recorded, treat this as 5 bowel movements -> current diarrhea]), OR
c) Stools described with a consistency that represents diarrhea, such as 'unformed', 'loose', 'watery', 'liquid', 'semi-formed', REGARDLESS OF FREQUENCY OF BOWEL MOVEMENTS.
- N.B: 'Soft' bowel movements are NOT considered to be diarrhea.
Edge cases:
- 'Clinical remission' - if the note states that a patient is in remission, then assume diarrhea is NOT present unless there is any other information in the note to suggest the presence of current diarrhea.
- 'Asymptomatic' or 'doing well' - if a note does not specifically mention the presence of diarrhea, but states the patient is asymptomatic or doing well, assume diarrhea is NOT present.
- If other gastrointestinal symptoms are documented in the note, but there is no mention of diarrhea, assume that diarrhea is NOT present.
- Contradictory information: some notes may contain contradictory information as to the presence/absence of diarrhea. If this is the case, use clinical judgement to decide whether the patient has current diarrhea as defined above.
###INSTRUCTIONS END###
Task: Use the above clinical note and the accompanying instructions to determine whether the patient has current diarrhea. Give an explanation and return one of the following labels: {'0: No current diarrhea', '1: Current diarrhea'}

*GitHub*

We have uploaded our annotation protocol, code for the tNLP models, and our finalized LLM prompts to GitHub (<https://github.com/rwelab/IBDNLP/tree/main>). We have also included instructions that allow other centers to customize these pipelines for their own institutions.

*Data Security*

All patient-identified data was stored and accessed in secure environments provided by the universities. All computers used for this project were encrypted by the university. We did not transfer patient data between institutions and instead uploaded the UCSF internally validated model to GitHub for download at Stanford University.

We used a HIPAA-compliant portal, UCSF Versa API interface, to interact with GPT-4. Through this secure portal, PHI data was sent to Azure OpenAI API, and we interacted with the LLM through a ChatGPT interface ^15,16^. Use of GPT-4 requires a secure interface with which PHI data can be sent to the Azure OpenAI API and therefore will be limited to UCSF data using the secure, HIPAA-compliant, UCSF Versa API interface. Similar HIPAA-compliant portals for GPT-4 and PaLM-2 were used at Stanford University.
