## Supplemental Tables for "Large language models outperform traditional natural language processing methods in extracting patient-reported outcomes in IBD"

| Task | Gender | Accuracy | Sensitivity | Specificity | Positive Predictive Value | Negative Predictive Value |
|---|---|---|---|---|---|---|
| Abdominal pain | Male (n=104) | 0.93 [0.89-0.97] | 0.96 [0.93-0.99] | 0.92 [0.88-0.96] | 0.85 [0.79-0.91] | 0.98 [0.96-1] |
| | Female (n=146) | 0.93 [0.88-0.98] | 0.96 [0.92-1] | 0.92 [0.87-0.97] | 0.82 [0.75-0.89] | 0.98 [0.96-1] |
| Diarrhea | Male (n=104) | 0.88 [0.83-0.93] | 0.91 [0.86-0.96] | 0.86 [0.80-0.92] | 0.84 [0.78-0.90] | 0.92 [0.88-0.96] |
| | Female (n=146) | 0.88 [0.82-0.94] | 0.87 [0.81-0.93] | 0.90 [0.84-0.96] | 0.87 [0.81-0.93] | 0.90 [0.84-0.96] |
| Fecal blood | Male (n=104) | 0.93 [0.89-0.97] | 0.88 [0.83-0.93] | 0.95 [0.91-0.99] | 0.88 [0.83-0.93] | 0.95 [0.91-0.99] |
| | Female (n=146) | 0.97 [0.94-1] | 1.00 | 0.96 [0.92-1] | 0.90 [0.84-0.96] | 1.00 |

*Supplemental Table 1: Bias assessment of PaLM-2 based on gender, conducted on the test set of 250 notes at Stanford University. Metrics are reported as percentages with their associated 95% confidence intervals.*

| Model | Disease | Accuracy | Sensitivity | Specificity | Positive Predictive Value | Negative Predictive Value |
|---|---|---|---|---|---|---|
| Abdominal pain | CD (n=138) | 0.93 [0.89-0.97] | 0.96 [0.93-0.99] | 0.91 [0.86-0.96] | 0.85 [0.79-0.91] | 0.98 [0.96-1] |
| | UC (n=112) | 0.94 [0.90-0.98] | 0.97 [0.94-1] | 0.93 [0.88-0.98] | 0.82 [0.75-0.89] | 0.99 [0.97-1] |
| Diarrhea | CD (n=138) | 0.91 [0.86-0.96] | 0.95 [0.91-0.99] | 0.88 [0.83-0.93] | 0.87 [0.81-0.93] | 0.96 [0.93-0.99] |
| | UC (n=112) | 0.85 [0.78-0.92] | 0.81 [0.74-0.88] | 0.88 [0.82-0.94] | 0.83 [0.76-0.90] | 0.86 [0.80-0.92] |
| Fecal blood | CD (n=138) | 0.94 [0.90-0.98] | 0.88 [0.83-0.93] | 0.96 [0.93-0.99] | 0.88 [0.83-0.93] | 0.96 [0.93-0.99] |
| | UC (n=112) | 0.96 [0.92-1] | 0.97 [0.94-1] | 0.95 [0.91-0.99] | 0.89 [0.83-0.95] | 0.99 [0.97-1] |

*Supplemental Table 2: Bias assessment of PaLM-2 based on diagnosis—Crohn's Disease (CD) versus Ulcerative colitis (UC). This was conducted on the test set of 250 notes at Stanford University. Metrics are reported as percentages with their associated 95% confidence intervals.*

| Task | Race | Accuracy | Sensitivity | Specificity | Positive Predictive Value | Negative Predictive Value |
|---|---|---|---|---|---|---|
| Abdominal pain | White (n=150) | 0.92 [0.88-0.96] | 0.95 [0.92-0.98] | 0.91 [0.86-0.96] | 0.78 [0.71-0.85] | 0.98 [0.96-1] |
| | Non-white (n=98) | 0.95 [0.91-0.99] | 0.97 [0.94-1] | 0.93 [0.88-0.98] | 0.90 [0.84-0.96] | 0.98 [0.95-1] |
| Diarrhea | White (n=150) | 0.91 [0.86-0.96] | 0.92 [0.88-0.96] | 0.90 [0.85-0.95] | 0.86 [0.80-0.92] | 0.94 [0.90-0.98] |
| | Non-white (n=98) | 0.85 [0.78-0.92] | 0.85 [0.78-0.92] | 0.84 [0.77-0.91] | 0.84 [0.77-0.91] | 0.86 [0.79-0.93] |
| Fecal blood | White (n=150) | 0.96 [0.93-0.99] | 0.94 [0.90-0.98] | 0.97 [0.94-1] | 0.89 [0.84-0.94] | 0.98 [0.96-1] |
| | Non-white (n=98) | 0.94 [0.89-0.99] | 0.91 [0.85-0.97] | 0.95 [0.91-0.99] | 0.91 [0.85-0.97] | 0.95 [0.91-0.99] |

*Supplemental Table 3: Bias assessment of PaLM-2 based on race conducted on the test set of notes at Stanford University. In our cohort, the category of Non-White race included Black (n=6), Pacific Islander (n=2), Native American (n=2), Asian (n=22), and Other (n=65). Two clinical encounters had an unknown race and were excluded. Metrics are reported as percentages with their associated 95% confidence intervals.*

| Task | Age Group | Accuracy | Sensitivity | Specificity | Positive Predictive Value | Negative Predictive Value |
|---|---|---|---|---|---|---|
| Abdominal pain | Below median (n=125) | 0.92 [0.87-0.97] | 0.94 [0.90-0.98] | 0.91 [0.86-0.96] | 0.81 [0.74-0.88] | 0.98 [0.96-1.00] |
| | Above median (n=125) | 0.93 [0.88-0.98] | 0.96 [0.92-1] | 0.92 [0.87-0.97] | 0.82 [0.75-0.89] | 0.98 [0.96-1.00] |
| Diarrhea | Below median (n=125) | 0.88 [0.83-0.93] | 0.91 [0.86-0.96] | 0.86 [0.80-0.92] | 0.84 [0.78-0.90] | 0.92 [0.88-0.96] |
| | Above median (n=125) | 0.88 [0.82-0.94] | 0.87 [0.81-0.93] | 0.90 [0.84-0.96] | 0.87 [0.81-0.93] | 0.90 [0.84-0.96] |
| Fecal blood | Below median (n=125) | 0.93 [0.89-0.97] | 0.88 [0.83-0.93] | 0.95 [0.91-0.99] | 0.88 [0.83-0.93] | 0.95 [0.91-0.99] |
| | Above median (n=125) | 0.97 [0.94-1] | 1.00 | 0.96 [0.92-1] | 0.90 [0.84-0.96] | 1.00 |

*Supplemental Table 4: Bias assessment of PaLM-2 based on age, conducted on the test set of 250 notes at Stanford University. We divided our cohort into above and below the median age of 35 years old. Metrics are reported as percentages with their associated 95% confidence intervals.*

| Task | Gender | Accuracy | Sensitivity | Specificity | Positive Predictive Value | Negative Predictive Value |
|---|---|---|---|---|---|---|
| Abdominal pain | Male (n=146) | 0.96 [0.93-0.99] | 0.90 [0.85-0.95] | 0.99 [0.97-1] | 0.98 [0.96-1] | 0.95 [0.91-0.99] |
| | Female (n=104) | 0.93 [0.88-0.98] | 0.86 [0.79-0.93] | 0.96 [0.92-1] | 0.89 [0.83-0.95] | 0.95 [0.91-0.99] |
| Diarrhea | Male (n=146) | 0.89 [0.84-0.94] | 0.80 [0.74-0.86] | 0.96 [0.93-0.99] | 0.95 [0.91-0.99] | 0.86 [0.80-0.92] |
| | Female (n=104) | 0.91 [0.85-0.97] | 0.80 [0.71-0.88] | 1 | 1 | 0.87 [0.81-0.93] |
| Fecal blood | Male (n=146) | 0.95 [0.91-0.99 | 0.88 [0.83-0.93] | 0.98 [0.96-1] | 0.95 [0.91-0.99] | 0.95 [0.91-0.99] |
| | Female (n=104) | 0.99 [0.97-1] | 0.96 [0.92-1] | 1 | 1 | 0.99 [0.97-1] |

*Supplemental Table 5: Bias assessment of GPT-4 based on gender, conducted on the test set of 250 notes at Stanford University. Metrics are reported as percentages with their associated 95% confidence intervals.*

| Model | Disease | Accuracy | Sensitivity | Specificity | Positive Predictive Value | Negative Predictive Value |
|---|---|---|---|---|---|---|
| Abdominal pain | CD (n=138) | 0.96 [0.93-0.99] | 0.89 [0.84-0.94] | 0.99 [0.97-1] | 0.98 [0.96-1] | 0.95 [0.91-0.99] |
| | UC (n=112) | 0.94 [0.90-0.98] | 0.86 [0.80-0.92] | 0.96 [0.92-1] | 0.89 [0.83-0.95] | 0.95 [0.91-0.99] |
| Diarrhea | CD (n=138) | 0.88 [0.83-0.93] | 0.77 [0.70-0.84] | 0.97 [0.94-1] | 0.96 [0.93-0.99] | 0.84 [0.78-0.90] |
| | UC (n=112) | 0.92 [0.87-0.97] | 0.83 [0.76-0.90] | 0.98 [0.95-1] | 0.98 [0.95-1] | 0.89 [0.83-0.95] |
| Fecal blood | CD (n=138) | 0.98 [0.96-1] | 0.91 [0.86-0.96] | 1 | 1 | 0.97 [0.94-1] |
| | UC (n=112) | 0.96 [0.92-1] | 0.91 [0.86-0.96] | 0.98 [0.95-1] | 0.94 [0.90-0.98] | 0.96 [0.92-1] |

*Supplemental Table 6: Bias assessment of GPT-4 based on diagnosis—Crohn's Disease (CD) versus Ulcerative colitis (UC). This was conducted on the test set of 250 notes at Stanford University. Metrics are reported as percentages with their associated 95% confidence intervals.*

| Task | Race | Accuracy | Sensitivity | Specificity | Positive Predictive Value | Negative Predictive Value |
|---|---|---|---|---|---|---|
| Abdominal pain | White (n=150) | 0.95 [0.92-0.98] | 0.87 [0.82-0.92] | 0.97 [0.94-1] | 0.92 [0.88-0.96] | 0.96 [0.93-0.99] |
| | Non-white (n=98) | 0.96 [0.92-1] | 0.92 [0.87-0.97] | 0.98 [0.95-1] | 0.97 [0.94-1] | 0.95 [0.91-0.99] |
| Diarrhea | White (n=150) | 0.91 [0.86-0.96] | 0.78 [0.71-0.85] | 0.98 [0.96-1] | 0.98 [0.96-1] | 0.87 [0.82-0.92] |
| | Non-white (n=98) | 0.89 [0.83-0.95] | 0.81 [0.73-0.89] | 0.96 [0.92-1] | 0.95 [0.91-0.99] | 0.84 [0.77-0.91] |
| Fecal blood | White (n=150) | 0.97 [0.94-1] | 0.91 [0.86-0.96] | 0.99 [0.97-1] | 0.97 [0.94-1] | 0.97 [0.94-1] |
| | Non-white (n=98) | 0.97 [0.94-1] | 0.90 [0.84-0.96] | 1 | 1 | 0.96 [0.92-1] |

*Supplemental Table 7: Bias assessment of GPT-4 based on race conducted on the test set of notes at Stanford University. In our cohort, the category of Non-White race included Black (n=6), Pacific Islander (n=2), Native American (n=2), Asian (n=22), and Other (n=65). Two clinical encounters had an unknown race and were excluded. Metrics are reported as percentages with their associated 95% confidence intervals.*

| Model | Age Group | Accuracy | Sensitivity | Specificity | Positive Predictive Value | Negative Predictive Value |
|---|---|---|---|---|---|---|
| Abdominal pain | Below median (n=125) | 0.94 [0.90-0.98] | 0.90 [0.85-0.95] | 0.96 [0.93-0.99] | 0.92 [0.88-0.97] | 0.95 [0.92-0.99] |
| | Above median (n=125) | 0.94 [0.90-0.98] | 0.86 [0.80-0.92] | 0.98 [0.96-1.00] | 0.97 [0.94-1.00] | 0.93 [0.88-0.97] |
| Diarrhea | Below median (n=125) | 0.90 [0.85-0.95] | 0.83 [0.76-0.90] | 0.97 [0.94-1.00] | 0.96 [0.93-0.99] | 0.87 [0.81-0.93] |
| | Above median (n=125) | 0.90 [0.85-0.95] | 0.77 [0.70-0.84] | 0.98 [0.96-1.00] | 0.98 [0.96-1.00] | 0.86 [0.80-0.92] |
| Fecal blood | Below median (n=125) | 0.97 [0.94-1.00] | 0.89 [0.84-0.94] | 1.00 | 1.00 | 0.96 [0.93-0.99] |
| | Above median (n=125) | 0.97 [0.94-1.00] | 0.93 [0.88-0.97] | 0.98 [0.96-1.00] | 0.93 [0.88-0.97] | 0.98 [0.96-1.00] |

*Supplemental Table 8: Bias assessment of GPT-4 based on age, conducted on the test set of 250 notes at Stanford University. We divided our cohort into above and below the median age of 35 years old. Metrics are reported as percentages with their associated 95% confidence intervals.*
